# Hypomethylation of Monoamine Oxidase A promoter/exon 1 region is associated with temper outbursts in Prader-Willi syndrome

**DOI:** 10.1101/2021.05.06.21256730

**Authors:** Maximilian Deest, Vanessa Buchholz, Kirsten Jahn, Christian Eberlein, Stefan Bleich, Helge Frieling

**Affiliations:** Department of Psychiatry, Social Psychiatry and Psychotherapy, Medical School Hannover, Hannover, Germany

**Keywords:** Prader-Willi syndrome, Monoamine Oxidase A, temper outbursts, Methylation

## Abstract

**Background:** Prader-Willi syndrome (PWS) is a rare neurodevelopmental disorder caused by the absence of paternally expressed and maternally imprinted genes on chromosome 15q 11.2-13. It is associated with a certain behavioural phenotype with repetitive and ritualistic behaviours, skin-picking and temper outbursts. Temper outbursts are characterized by verbal and physical aggression with screaming, destroying property, and/or physical aggression towards others. They drastically effect the quality of life of the individuals as well as the relatives and caregivers. Recent studies show a promising therapeutic effect of serotonin reuptake inhibitors like sertraline on frequency and intensity of outbursts. Monoamine oxidase A (MAOA) (X p11.23) plays a crucial role in the metabolism of monoamines such as serotonin, norepinephrine, and dopamine. Dysregulation in methylation of the CpG island spanning the promoter region and exon 1 of MAOA is implicated in impulsive and aggressive behaviour.

**Methods:** In the present study, methylation rates of CpG dinucleotides in the MAOA promoter and exon 1 region were determined from DNA derived from whole blood samples of PWS patients (n=32) and controls (n=14) matched for age, sex and BMI via bisulfite sequencing. PWS patients were grouped into those showing temper outbursts, and those who do not.

**Results:** Overall, PWS patients show a significant lower methylation rate at the promoter/exon 1 region than healthy controls in both sexes. Furthermore, PWS patients, male as well female with temper outbursts show a significant lower methylation rate than those without temper outbursts (p<0.001 and p=0.006)

**Conclusion:** The *MAOA* promoter/exon 1 region methylation seems to be dysregulated in PWS patients in sense of a hypomethylation, especially in those suffering from temper outbursts. As *MAOA* is involved in the metabolism of serotonin, we conclude that this dysregulation plays a crucial role in the pathophysiology of temper outbursts in PWS. Furthermore, our findings suggest, that dysregulation of certain genes outside the PWS locus contribute to the behavioural phenotype of PWS.

## 1. Introduction

Prader-Willi syndrome (PWS) is a rare neurodevelopmental disorder associated with a certain behavioural phenotype with repetitive and ritualistic behaviours, skin-picking, and temper outbursts (Sinnema *et al*., 2011; Whittington and Holland, 2018; Butler, Miller and Forster, 2019). These temper outbursts are characterized by verbal and physical aggression with screaming, destroying property, and/or physical aggression towards others (punching, kicking etc) (Rice, Woodcock and Einfeld, 2018a).

PWS, in general, is characterised by hypogonadism and growth hormone deficiency, mild to moderate intellectual disability, and infantile hypotonia with an early failure to thrive. Newborns with PWS typically display feeding difficulties that drastically changes during early childhood with the development of excessive hyperphagia leading to severe obesity, if food intake is not restricted (Dykens and Shah, 2003; Cassidy *et al*., 2012; Angulo, Butler and Cataletto, 2015). The cause of PWS is the lack of expression of paternally expressed but maternally imprinted genes on the chromosome 15q11.2 – 13 region. This can either result from a paternal deletion (delPWS; 70% of cases), maternal uniparental disomy (mUPD;25%), or an imprinting centre defect (IC;3-5%) (Nicholls and Knepper, 2001; Horsthemke and Wagstaff, 2008). Several mental disorders, such as autism spectrum disorders (ASD), psychosis, and depression, are more frequent in PWS than in other populations. The prevalence of mental disorders differs among genetic subtypes of PWS; with a higher rate of ASD and psychosis in those with a mUPD than those with a delPWS (Holland *et al*., 2003; Vogels *et al*., 2003; Soni *et al*., 2007, 2008).

Temper outbursts are displayed by up to 88% of individuals with PWS (Dykens, Cassidy and King, 1999), typically start in childhood and increase in intensity and frequency with age (Dykens *et al*., 1992). They drastically affect the quality of life of the individual as well as the relatives and caregivers (Mazaheri *et al*., 2013). Recent studies identified the provocation of those temper outbursts as goal blockage, difficulty in dealing with change, and feeling of social injustice (Rice, Woodcock and Einfeld, 2018a). Current treatment options include clinical behavior interventions like token boards or behavior management plans. Several studies have shown that the serotonergic system is disturbed in PWS, possibly contributing to the behavioural phenotype. For instance, SNORD115 (HB-52 III small nuclear RNA), located at the PWS region is involved in alternative splicing of the serotonergic 2c receptor (Kishore and Stamm, 2006), and mice with a deletion of the IC show reduced functioning of the serotonin 2c receptor. Those mice showed increased impulsivity and compulsivity. The use of a serotonin receptor agonist improved impulsivity in these mice (Davies *et al*., 2019). Accordningly, we recently showed that the selective serotonin reuptake inhibitor (SSRI) sertraline is a promising treatment option for temper outbursts in PWS leading to a decrease of frequency and intensity of outbursts (Deest *et al*., 2020). All in all, these findings strongly suggest that the serotonergic system may play a role in the pathophysiology of behaviour problems in PWS.

The *monoamine oxidase A gene*, located on Xp11.4-p11.3, and its product MAOA play a crucial role in the metabolism of monoamines such as serotonin, norepinephrine, and dopamine (Lan *et al*., 1989; Bortolato, Chen and Shih, 2008). *MAOA* consists of 14 exons and 14 introns with two cytosine-guanine (CpG) islands. One CpG island is located at the promoter region and the second one spans the promoter region as well as exon I and the beginning of intron I (Shih, Wu and Chen, 2011). Previous studies showed significant disturbances in methylation of these CpG islands in several mental disorders like anxiety disorders (Domschke *et al*., 2012), depression (Melas *et al*., 2013; Melas and Forsell, 2015), posttraumatic stress disorder (PTSD) (Ziegler *et al*., 2018) and antisocial personality disorder/conduct disorder (Cecil *et al*., 2018). Individuals with Brunner syndrome, caused by a MAOA deficiency, show serotonergic symptoms (flushing, diarrhea, headache, palpitations) and aggressive outbursts (Palmer *et al*., 2016). In PWS, Akefeldt et al. showed, that MAO activity in platelets is significantly higher than in controls (Åkefeldt and Månsson, 1998).

As disturbances of the serotonergic pathway most likely contribute to temper outbursts and *MAOA* is crucially involved in the metabolism of serotonin, we here investigate the methylation of *MAOA* in PWS. Therefore, we investigated the methylation of 47 CpG nucleotides at the CpG island spanning exon 1 and part of the promoter to investigate methylation between PWS and healthy individuals with a focus on temper outbursts.

## 2. Methods

### 2.1 Subjects

This study was approved by the local Ethics Committee of Hannover Medical School and adhered to the Declaration of Helsinki. All participants in this study gave their written informed consent for participation in this study and were recruited at the Outpatient Department for Mental Health in Rare Genetic Disorders of the Department of Psychiatry, Social Psychiatry, and Psychotherapy of Medical School Hannover, Hannover, Germany. All participants were patients of the Outpatient Department. In patients, the PWS diagnosis was genetically confirmed. Characteristics such as age, gender, and BMI were documented. The controls were matched for gender, body mass index (BMI), and age. EDTA-blood was collected and stored at Hannover Unified Biobank.

### 2.2 Assessment of temper outbursts

As currently no reliable psychometric test instrument in german language for temper outbursts in PWS exist, the caregivers of the patients had to state if temper outbursts take place and have a significant impact on the patients daily life. A criterion for this was that the patient must exhibit physical aggression towards others and/or destroy property.

### 2.3 DNA isolation and bisulfite reaction

DNA for methylation analysis was extracted from blood directly after sampling by the Hannover Unified Biobank using the Hamilton ChemagicStar (Hamilton Germany Robotics, Graefelfing, Germany) and the chemagicStar DNA-Blood1k kit (PerkinElmer chemagen Technology, Baesweiler, Germany). Bisulfite conversion and purification was performed using the EpiTect® 96 Bisulfite Kit (Qiagen, Hilden, Germany) following the manufacturer recommendations.

### 2.4 Amplification of target sequences

Amplification of *MAOA* promoter, exon 1/ intron 1 target sequences of the purified bisulfite- converted DNA was conducted using touchdown PCRs. We used the forward primer ‘GTTATAAAGGGGTTAGATTAAGTGG’ and the reverse primer ‘AAACCTAAACAATTATACCCCC’. Cycling conditions apply as follows: 15 min 95 °C, 1 min 97 °C, 15 x (30s 95 °C, 45s 65 °C with −1 °C per cycle, 1 min 72 °C), 30 x (30s 95°C, 45s 50 °C, 1 min 70 °C), 5 min 65 °C. Primers were ordered from Integrated DNA Technologies (Coral Ville, IA, USA). PCRs were performed on a C1000 Thermal Cycler (BIO-RAD, Hercules, CA, USA) using the HotStarTaq® Master Mix Kit (QIAGEN, Hilden, Germany). The PCR products were purified automatically on a Biomek® NxP (Beckman Coulter, Brea, CA, USA) using paramagnetic beads (Clean-NGS, GC Biotech®).

### 2.5 Sequencing

The Big-Dye® Terminator v3.1 Cycle Sequencing Kit (Applied Biosystems, Foster City, CA, USA) was used for sequencing PCR of the target fragments. Oligonucleotides used for sequencing PCR were ‘GTTATAAAGGGGTTAGATTAAGTGG’. Cycling conditions apply as follows: 1 min 96°C, 25 x (5s 96°C, 90s 60°C, 90s 50°C). Clean-up of the sequencing PCR was also automatized on our Biomek Nxp by use of CleanDTR beads (CG Biotech®). Sequencing was performed on a 3500XL genetic analyzer from ABI Life Technologies (Grand Island, NY, USA) according to the manufacturer’s instructions. The amplified fragments covered 47 CpG sites within exon 1 and the promoter region of the *MAOA* gene (from −241 bp to +328 bp of the first exon). Position of CpG sites is given in relation to the starting point of exon 1: GRCh38:X:43654907 (from ENSEMBL#ENSG00000189221).

### 2.6 Determination of methylation Rates

Determination of methylation rates for each CpG site and sequence analysis were performed using the Epigenetic Sequencing Methylation analysis software (ESME) (Lewin *et al*., 2004). The ratio between normalized peak values of cytosine and thymine was used to calculate the methylation rate of each CpG site per subject.

### 2.7 Statistical analysis

Sequence quality was determined via Sequence Scanner v2.0 software (ABI Life Technologies). All sequences showed a Quality Value > 20 for Trace Score in the Quality Control Report and could therefore be included for further analyses. All statistical analysis were performed using the Statistical Package for Social Sciences (SPSS, IBM, Armonk, NY). GraphPad Prism was used for data illustration. Single CpGs with less than 95% sequencing success among the samples were excluded which applied for three CpGs ahead of the first exon as well as twenty of the first exon. All samples showed more than 95% valid measurements and thus had been included. No CpG site showed less than 5% inter-individual variability. After applying these critera for data exlusion, the number of analyzed CpGs was 24. Mixed linear models were calculated to detect significant fixed effects of different variables on methylation rate. Bonferroni correction method was used to correct P values for multiple testing. In each analysis, a p value of < 0.05 was considered significant.

## 3. Results

### 3.1 Demographic data

In this study we included 32 patients with genetically confirmed diagnosis of PWS (13 female, 19 male) with a mean age of 27.9 years (min: 12y, max:55y) and a mean BMI of 28.1 kg/m^2^ (min: 19.1 kg/m^2^, max: 55.58 kg/m^2^). The control group consisted of 14 healthy individuals without PWS (5 female, 9 male) with a mean age of 31.2 years (min: 18y, max:47y) and a mean BMI of 26.2 kg/m^2^ (min: 19.4 kg/m^2^, max: 40.1 kg/m^2^) (see Table 1 for characteristics of study population). In 15 cases PWS was caused by a deletion (46.9%), with four having a deletion type I, 10 having a deletion type II. In one case no differentiation between deletion subtype was performed during genetic counseling. In nine patients PWS was caused by a mUPD (28.1%), in 4 cases by IC (9.4%). In four cases distinguishing between mUPD or IC did not take place due to various reasons (e.g. no blood from the parents could be obtained).

**Table 1.** Demographics of study population. Study population is groubed by controls and PWS. PWS patients are then grouped by showing temper outbursts vs. not showing outbursts. Abbrevations: del, deletion; del I, deletion type I; del II, deletion tye II, UPD maternal uniparental disomie; IC, Imprinting defect; OCD, obsessive-compulsive disorder; ASD, autism spectrum disease, ns, not specified because no differentiation of specific subtype could be obtained (del I vs del II or UPD vs IC) due to different reasons (e.g.) no blood from the parents could be obtained

|  | control (n=14) | PWS |  |  |
| --- | --- | --- | --- | --- |
|  |  | total (n=32) | with temper outbursts (n=17) | no temper outbursts (n=15) |
| Sex | 9 male (64.3%)<br>5 female (35.7%) | 19 male (59.4%)<br>13 female (40.6%) | 11 male (64.7%)<br>6 female (35.3%) | 8 male (53.3%)<br>7 female (46.7%) |
| Age | 18-47 y (mean 31.21y) | 12-55y (mean 27.9y) | 12-41y (mean 24.8y) | 15 - 55y (mean 29.3y) |
| BMI | 19.4-40 kg/m2 (mean 26.2 kg/m2) | 19.1 - 55.1 kg/m2 (mean 28.1 kg/m2) | 19.1-55.6kg/m2 (mean 29kg/m2) | 19.9 - 36.5y (mean 26.96kg/m2) |
| Subtyp | / | del: 15 (4 del I, 10 del II, 1 ns)<br>UPD/IC: 17 (9 UPD, 4 IC, 4 ns) | del: 9 (2 del I, 6 del II, 1 ns)<br>UPD/IC: 8 (5 UPD, 2 IC, 1 ns) | del: 6 (2 del I, 4 del II)<br>UPD/IC: 9 (4 UPD, 2 IC, 3 ns) |
| Antipsychotic medication | / | n= 18 (56%) | n = 8 (44%) | n=8 (57%) |
| Antidepressant medication | / | n=4 (12.5%) | n=1 (5.5%) | n=3 (21.4%) |
| Psychiatric diagnosis | / | psychosis = 7<br>OCD = 2<br>ASD = 2 | psychosis = 2<br>ASD = 2 | psychosis=5<br>OCD=2 |

At the timepoint of counseling, caregivers had to state (yes/no) if temper outbursts are common and have a significant impact on the daily life. Of the 32 individuals, 17 (eleven male, six female) showed temper outbursts and 15 (eight male, seven female) did not (see Table 1 for more characteristics).

Overall, 18 patients (56%) received an antipsychotic medication (e.g. risperidone, aripiprazole, quetiapine) and four received an antidepressant medication (e.g. sertraline, fluoxetine) at timepoint of consultation. Seven patients suffered from a psychosis (acute or chronic), two from an obsessive-compulsive disorder and two from a ASD. The other patients received medication because of behavioural problems. In the subgroup of those showing temper outbursts eight (44%) received an antipsychotic medication and one (5.5%) an antidepressant. In those showing no outbursts, eight (57%) received antipsychotic medication and three (21.4%) received an antidepressant medication.

### 3.2 The difference in Methylation Rate between PWS and controls

Comparison of the mean methylation levels of the investigated CpGs within the *MAOA* exon 1/promoter region showed significantly lower methylation rates in PWS than in healthy controls (p<0.001) with a mean methylation of 17.8% +/-1% for controls and 11.8% +/-0.6% for PWS. (see Fig. 1A). Methylation levels are known to be gender-dependent in general (Singmann *et al*., 2015) and for *MAOA* in particular (Chen *et al*., 2012; Domschke *et al*., 2012), so we performed a gender-specific analysis. Looking at the mean methylation rate in males, PWS subjects showed a mean methylation rate of 1.1% +/- 0.4%, and male controls had a value of 6.9% +/- 0.6%. The difference between those two groups was significant (p<0.001) (see Fig. 1B). In the female PWS group mean methylation rate was 26.3% +/- 0.9% and in the control group the value was 37.2% +/- 1.4%. The difference in methylation rate between female PWS subjects and female controls was also significant (p<0.001) (see Fig. 1C). As mean methylation rates showed a strong differences between the sexes, we performed only gender specific analysis for the next steps.

**Figure 1.**
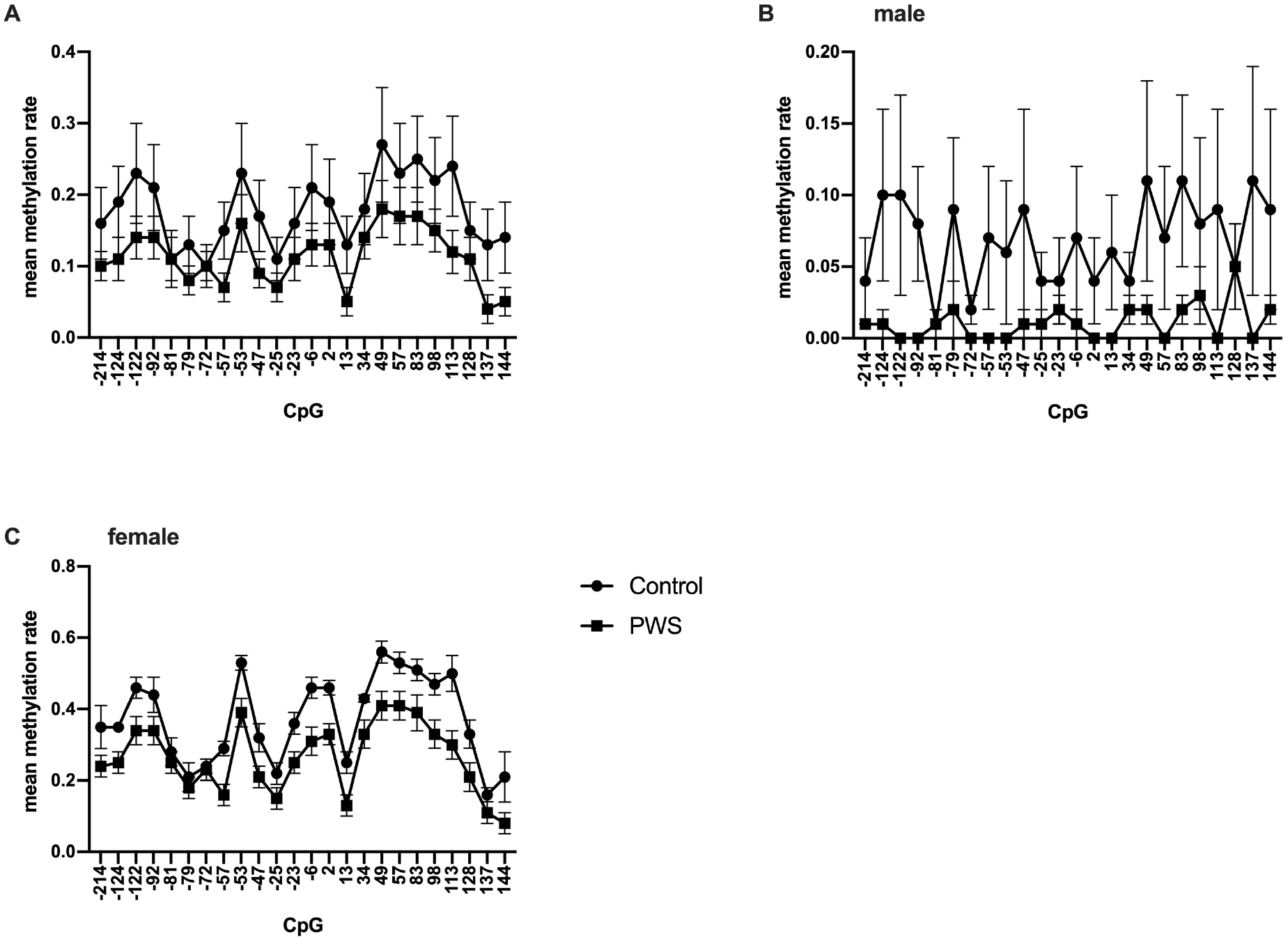
Overview of methylation rates for single CpGs. (A) PWS patients show a significant lower methylation than controls, (B), (C) Methylation rates between PWS and controls in regard of sex show, that methylation in PWS is significantly lower than in healthy controls in both sexes

### 3.3 The difference in methylation rate between the genetic subtype of PWS

As there are certain differences, especially in the distribution of psychiatric diseases between the genotypes of PWS, we were interested in differences in methylation between those groups. As an IC and a mUPD lead to the same imprinting pattern in the PWS region (Horsthemke and Wagstaff, 2008), we split our cohort into two groups: a delPWS subgroup and a mUPD/IC subgroup. This resulted in groups of 15 individuals with a delPWS (ten male, five female) and 17 with a mUPD/IC (nine male, eight female). When looking at the males, those with the delPWS subtype showed a mean methylation rate of 2.1% +/- 0.6% compared with those with an mUPD/IC with a mean methylation rate of 0% +/- 0.7%. Those with a deletion subtype showed significantly lower methylation rates (p<0.001) compared to healthy individuals as well as those with an mUPD/IC (p<0.001). The difference between the subtypes was not significant (p=0.053) (see Fig. 2A). In the female group, the mean methylation rate of those with a delPWS was 28.4% +/- 1.4% and those with mUPD/IC was 24.9% +/- 1.1%. Both delPWS as well mUPD/IC groups were significantly different (p<0.001) from healthy controls. However, there was no significant difference between the genetic subgroups (p=0.147) (see Fig. 2B).

**Figure 2.**
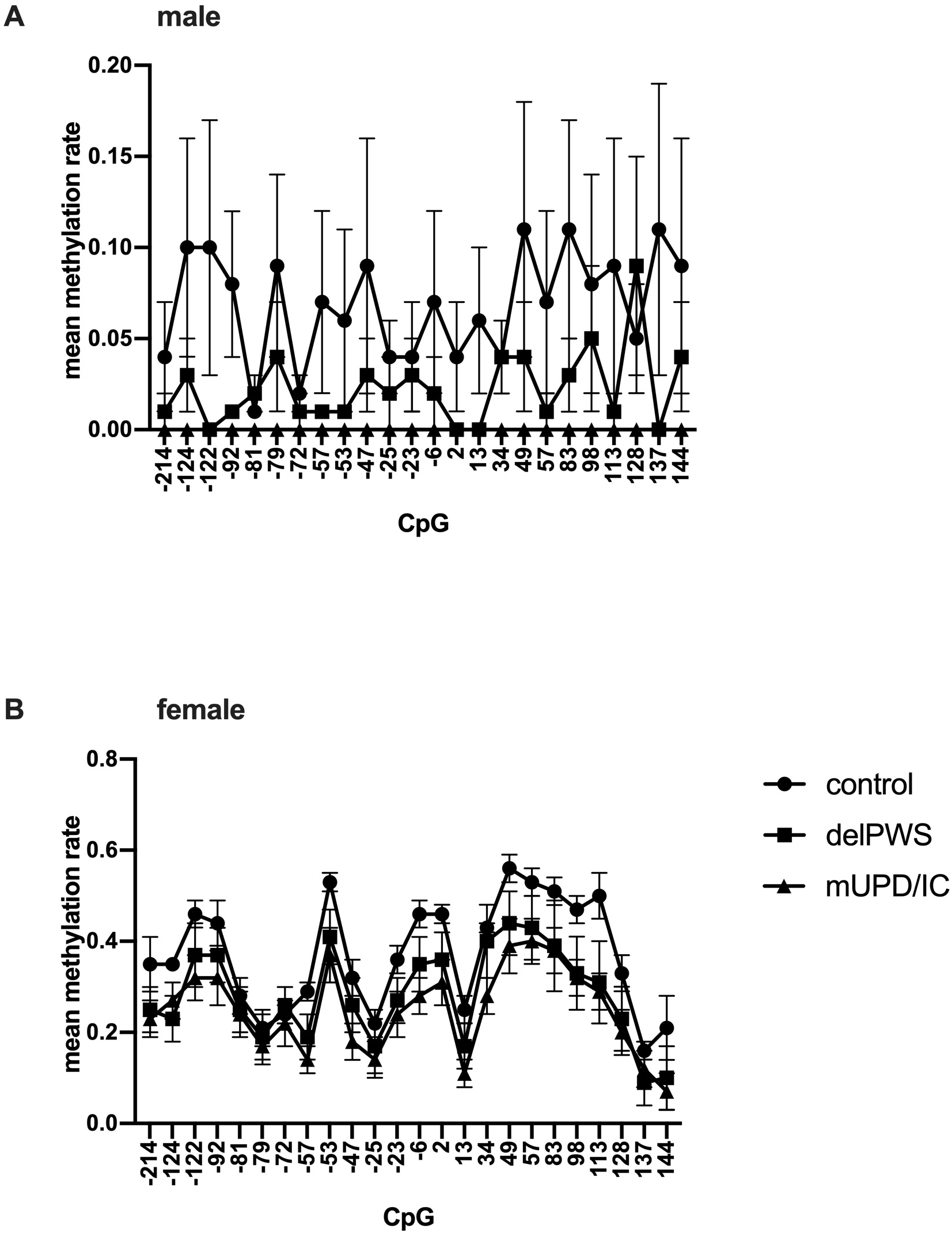
Comparison of methylation rates between genetic subtypes of PWS and healthy controls in male (A) and female (B). No difference between the subtypes was observed.

### 3.4 Temper outbursts and Methylation Rate

As temper outbursts are common among all genetic subtypes we next investigated the methylation rate among those showing temper outbursts and those not showing temper outbursts. The caregivers had been asked if temper outbursts are common and have a significant impact on daily life. Of the 32 individuals with PWS in this study, 17 showed temper outbursts (11 male, six female) and 14 did not (eight male, six female).

We again compared the mean methylation rate in a sex-specific manner. In healthy males the mean methylation rate was 6.9% +/- 0.6%, in male PWS showing no temper outbursts the value was 2.7% +/- 0.7% and in those with temper outbursts 0% +/- 0.6%. Thus male PWS without temper outbursts shows significantly lower methylation than healthy male controls (p<0.001) but significantly higher methylation than those PWS individuals with temper outbursts (p=0.006) (see Fig. 3A).

**Figure 3.**
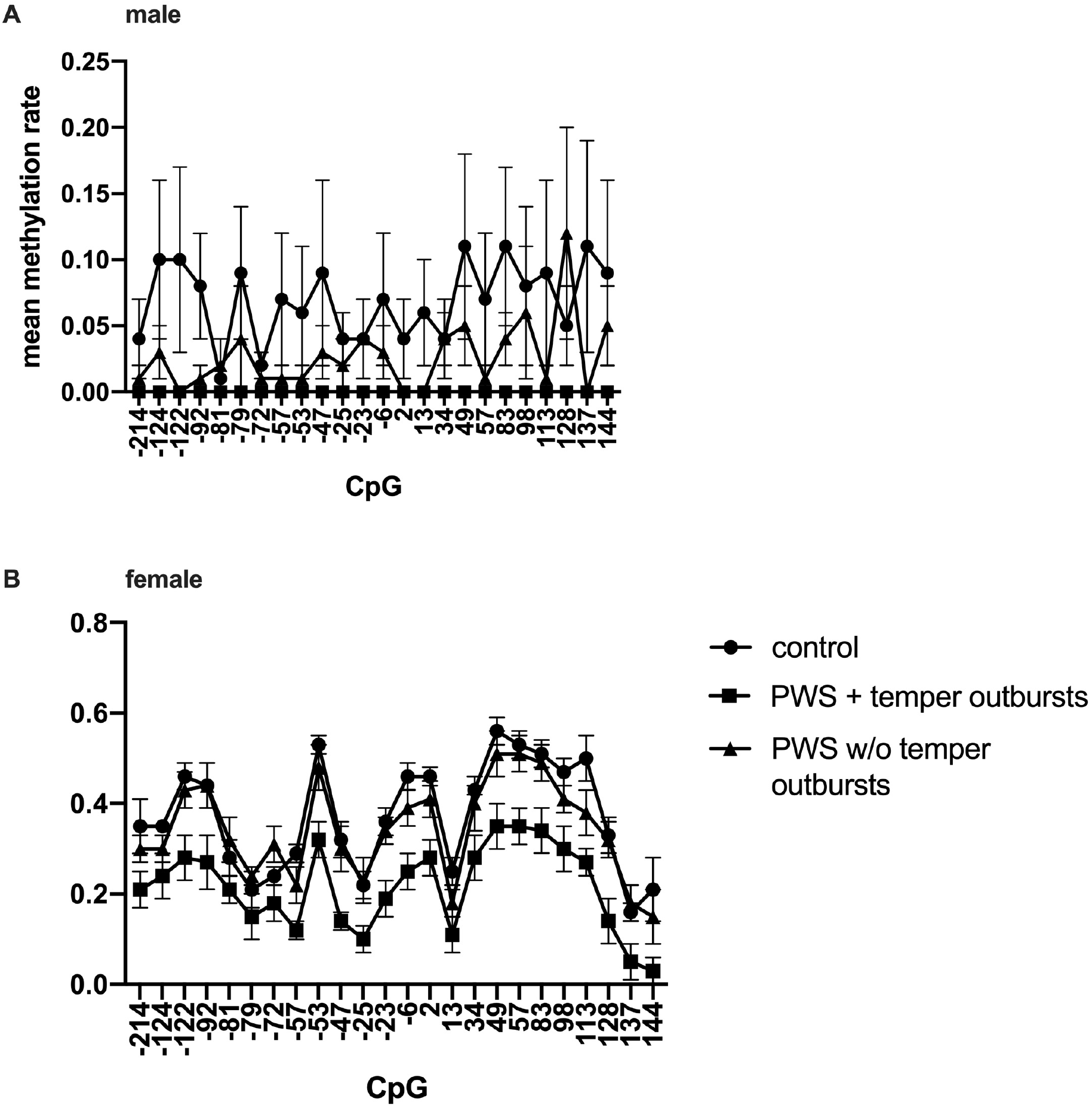
Comparison of methylation rates between PWS individuals showing temper outbursts, PWS individuals not showing temper outbursts and healthy controls in male (A) and female (B). In both sexes, patients suffering from temper outbursts have a significant hypomethylation than healthy controls and PWS patients without outbursts.

In female healthy controls the mean methylation rate was 37.2% +/- 1.4%, in female PWS without temper outbursts the value was 30.4% +/- 1.1% and in female PWS showing temper outbursts the value was 21.4% +/- 1.2 resulting in no significant difference between healthy controls and female PWS showing no temper outbursts (p=0.265). However, females with PWS showing temper outbursts had significantly lower methylation than healthy controls (p<0.001) and PWS individuals without temper outbursts (p<0.001) (see Fig. 3B).

## 4. Discussion

In this study, we investigated the methylation pattern of the CpG island flanking exon 1 and part of the promoter region in *MAOA* in PWS in comparison with healthy controls matched for age, sex, and BMI. In our study group, male controls as well as male PWS individuals, show significantly lower methylation than females which is in line with previous findings (Domschke *et al*., 2012; Singmann *et al*., 2015). However, in both sexes, the methylation rate in PWS is significantly lower than in healthy controls. There are several reported findings of differences in the prevalence of certain psychiatric disorders, such as psychosis or ASD, between genetic subtypes of PWS (Vogels *et al*., 2003; Soni *et al*., 2007, 2008; Dykens *et al*., 2017). However, we did not find a significant difference in methylation rates between delPWS or mUPD/IC. Thus differences in *MAOA* methylation probably do not contribute to the described differences between the subtypes. Temper outbursts, often associated with physical aggressiveness and/or self-harm are a major problem across all genetic subtypes of PWS (Manzardo *et al*., 2018; Rice, Woodcock and Einfeld, 2018b). In our study cohort, 53% showed frequent temper outbursts having a significant impact on daily life as rated by the caregivers. As dysregulation of methylation in exon 1 of *MAOA* is known to be associated with impulsiveness and aggression (Cecil *et al*., 2018), we compared the methylation patterns of PWS individuals with temper outbursts and those without. In both sexes, individuals with temper outbursts showed a significantly lower mean methylation rate than those without temper outbursts and healthy controls. As the rate of those receiving an antipsychotic or antidepressant medication was similar between those groups, we think that the observed difference is not caused by the medication.

Methylation can be tissue-specific and as we investigated the methylation rate using peripheral blood, it could be argued that this might not predict central enzyme activity. However previous results showed that peripheral methylation of *MAOA* correlates with the central enzyme activity of MAOA using in vivo brain imaging via positron emission tomography (Shumay *et al*., 2012). Furthermore, it has been shown that methylation of the first exon predicts protein expression in general (Brenet *et al*., 2011).

Thus, we conclude that individuals with PWS, especially those with temper outbursts, have an increased MAOA activity in the brain leading to increased metabolism of monoamines, especially serotonin. It has been shown that platelet MAO activity, both very low and very high, correlates with impulsivity in children but not with intellectual levels (Shekim *et al*., 1984). This would be in line with previous findings from Akesfeldt et al. which showed increased serotonin metabolites in cerebrospinal fluid in PWS indicating an increased metabolism of serotonin (Åkefeldt *et al*., 1998). We previously showed that PWS patients having temper outbursts benefitted from medication with the SSRI sertraline (Deest *et al*., 2020). This inhibits the re-uptake of serotonin and thus increases the intrasynaptic serotonin concentration and thus counteracts the increased MAOA activity. Disturbances of the serotonergic system have long been proposed to contribute to the behaviour problems of PWS. One hypothesis is, that *SNORD 115* (HBII-52), a small nuclear RNA located at the PWS IC regulates alternative splicing of the serotonin 2c receptor (Kishore and Stamm, 2006; Davies *et al*., 2019). However, Hebras et al. recently showed that in knockout *SNORD115* mice alternative RNA splicing is unchanged and mice did not show significant behaviour aberrations. They raised doubts about the involvement of alternative RNA splicing of serotonin 2c receptor in the pathophysiology of behavioural problems (Jade *et al*., 2020). Our findings could lead to the hypothesis that it is not the dysregulation of serotonin receptors but dysregulation of MAOA activity that contributes to the disturbance of the serotonergic system in PWS. This could be in line with findings that mouse models of PWS showing aberrant behaviour have decreased impulsivity when treated with a serotonin receptor agonist (Davies *et al*., 2019). Interestingly, MAOA inhibitors are treatment options for atypical depression that is usually associated with symptoms like overeating and sleep increase, which are particular key symptoms of PWS (Davidson *et al*., 1982).

Our findings could also contribute to the understanding of temper outbursts in PWS in general. Several mental disorders are associated with disruptions of methylation patterns in *MAOA*. Two studies showed lower methylation at the CpG sites in *MAOA* promoter as well as the exon I /intron I region in female patients with an anxiety disorder (Domschke *et al*., 2012; Ziegler *et al*., 2016), as well as in Acrophobia (Schiele *et al*., 2018). Furthermore, several studies showed lower methylation rates in female patients with depression (Melas *et al*., 2013) and mixed anxiety depression and dysthymia (Melas and Forsell, 2015). It was also shown that lower methylation was associated with a higher score in Beck`s Depression Inventory (Peng *et al*., 2018). On the other hand, it was shown that higher methylation rates are found in conduct disorder (Cecil *et al*., 2018), Borderline personality disorder (Dammann *et al*., 2011), PSTD (Ziegler *et al*., 2018) and schizophrenia (Chen *et al*., 2012). It has been shown that temper outbursts in PWS occur more frequently during cognitive challenges or at an unexpected change of routines resulting in a cognitive overload with a loss of control (Woodcock, Oliver and Humphreys, 2009; Woodcock *et al*., 2010). Manning et al. showed that transcutaneous vagus nerve stimulation is a promising treatment option for temper outbursts (Manning *et al*., 2019). They propose that the rigidity in thinking leads to a flight or fight response to feeling under threat associated with increased anxiety. This rising anxiety consequently serves as the motor for the outbursts. We think that our findings, which are in line with those in patients with anxiety or depression, support this hypothesis. Furthermore, our data support the hypothesis that impulsiveness in PWS should not be seen in a link with mental disorders such as conduct disorder or personality disorders.

## 5. Conclusion

In this study we investigated the methylation pattern of exon 1 and part of the promoter region of *MAOA* in PWS. Overall, individuals in PWS who show temper outbursts have a significantly lower methylation rate at the investigated CpGs than those PWS individuals who do not have temper outbursts and healthy controls. This leads us to the conclusion that *MAOA* is critically involved in the disturbance of the serotonergic pathway in PWS and contributes to the high frequency of temper outbursts.

## Data Availability

Data available on request due to privacy/ethical restrictions

## Acknowledgement

We thank Prof. Anthony J. Holland, University of Cambridge, for his expertise and comments that greatly improved the manuscript.

